# Adapting Biomedical Foundation Models for Predicting Outcomes of Anti Seizure Medications

**DOI:** 10.1101/2025.08.07.25333198

**Authors:** Duy Khoa Pham, Deval Mehta, Yiwen Jiang, Daniel Thom, Richard Shek-kwan Chang, Mohammad Nazem-Zadeh, Emma Foster, Timothy Fazio, Sarah Holper, Karin Verspoor, Jiahe Liu, Duong Nhu, Sarah Barnard, Terence O’Brien, Zhibin Chen, Jacqueline French, Patrick Kwan, Zongyuan Ge

**Affiliations:** AIM for Health Lab, Faculty of IT, Monash University, Melbourne, Australia; Faculty of Engineering, Monash University, Melbourne, Australia; School of Translational Medicine, Monash University, Melbourne, Australia; Clinical Informatics Centre, Royal Melbourne Hospital, Melbourne, Australia; Department of Medicine, University of Melbourne, Melbourne, Australia; Royal Melbourne Hospital, Melbourne, Australia; School of Computing Technologies, RMIT University, Melbourne, Australia; NYU Langone Health, New York City, New York, United States; Department of Computing Technologies, Swinburne University of Technology, Melbourne, Australia

**Keywords:** instruction tuning, epilepsy, drug recommendation

## Abstract

Epilepsy affects over 50 million people worldwide, with anti-seizure medications (ASMs) as the primary treatment for seizure control. However, ASM selection remains a “trial and error” process due to the lack of reliable predictors of effectiveness and tolerability. While machine learning approaches have been explored, existing models are limited to predicting outcomes only for ASMs encountered during training and have not leveraged recent biomedical foundation models for this task. This work investigates ASM outcome prediction using only patient MRI scans and reports. Specifically, we leverage biomedical vision-language foundation models and introduce a novel contextualized instruction-tuning framework that integrates expert-built knowledge trees of MRI entities to enhance their performance. Additionally, by training only on the four most commonly prescribed ASMs, our framework enables generalization to predicting outcomes and effectiveness for unseen ASMs not present during training. We evaluate our instruction-tuning framework on two retrospective epilepsy patient datasets, achieving an average AUC of **71.39** and **63.03** in predicting outcomes for four primary ASMs and three completely unseen ASMs, respectively. Our approach improves the AUC by **5.53** and **3.51** compared to standard report-based instruction tuning for seen and unseen ASMs, respectively. Our code, MRI knowledge tree, prompting templates, and TREE-TUNE generated instruction–answer tuning dataset are available at the [link](https://github.com/khoapham154/Knowledge-Tree-Driven-Contextualized-Instruction-Tuning-of-Foundation-Models).

## 1 Introduction

Epilepsy is one of the most common neurological disorders, affecting over 50 million people worldwide [28]. Antiseizure medications (ASMs) are the first-line treatment for newly diagnosed patients, aiming to achieve seizure freedom, yet only about half of newly diagnosed patients achieve seizure control with their initial ASM [3]. ASM selection primarily depends on epilepsy type, clinical history and diagnostic findings. However, since ASMs exhibit similar efficacy for specific epilepsy types [20], ASM selection often follows a “trial and error” approach, requiring sequential trials of different ASMs if seizures persist, which results in multiple ineffective treatments before identifying the right ASM [4].

The epilepsy research community has explored machine learning (ML) and deep learning (DL) for ASM recommendation by identifying patterns linking patient health data to ASM outcomes [5]. Early studies used simple ML models such as kNN and random forests to predict the response to ASM based on genetic data [22, 24]. Recent works have incorporated the clinical history, demographics, and risk factors of patients, employing SVM, XGBoost, and MLP to predict their ASM outcomes [6,12]. More recently, researchers have tapped into applying ML for analyzing routine electroencephalogram (EEG) data to predict the most effective ASM to achieve seizure free outcome [8, 23].

The International League Against Epilepsy recommends the use of Magnetic Resonance Imaging (MRI) for newly diagnosed epilepsy patients, as it can detect structural lesions that can guide treatment decisions and epilepsy management. Early MRI findings help clinicians determine whether to escalate ASM trials or consider adjunct therapies like lesion resection [14]. Lesions like focal cortical dysplasia and mesiotemporal sclerosis have well-documented links to clinical outcomes and recognized patterns of drug responsiveness or resistance [1]. Research on MRI-based ASM outcome prediction has primarily relied on manually extracted features and simple ML models or CNNs, limiting their predictive power [13, 15, 26]. Moreover, existing studies operate in a closed-set setting, simply encoding ASMs as categorical input variables during training, which limits their ability to predict outcomes for unseen ASMs.

To the best of our knowledge, no prior work has explored recent biomedical foundation models [16, 30] for ASM outcome prediction. These models, trained on large-scale biomedical images and text, have demonstrated strong medical context understanding. However, they have struggled with custom and unseen downstream tasks, diverse demographics, and open-set questions [21, 29]. To address this, the broader vision and natural image research communities have adopted instruction tuning [2,17] as an efficient fine-tuning approach to improve model performance for custom downstream tasks.

### Our contributions

In this work, we explore ASM outcome prediction using MRI scans and reports. Our main contributions are: 1) **Foundation Model Adaptation**- We adapt biomedical vision-language foundation models to assess their efficacy for the task of ASM outcome prediction; 2) **Knowledge Tree Driven Contextualized Instruction Tuning (TREE-TUNE)**- To enhance the performance of foundation models, we construct a contextualized instruction-tuning dataset. Using an expert-built MRI knowledge tree, we categorize MRI reports and scans, generating data-specific instruction-answer pairs with GPT-4o for improved fine-tuning; 3) **Molecular Feature Integration**-To enable outcome predictions for unseen ASMs, we encode ASMs as Simplified Molecular Input Line Entry System (SMILES) [27] representations, enabling models to learn molecular features instead of treating ASMs as categorical variables.

We benchmark our approach on two retrospective epilepsy datasets, achieving a **5.53** AUC improvement over standard text-based instruction tuning for four primary ASMs. Notably, our model attains **63.03** AUC for predicting outcomes of three unseen ASMs it has never encountered during training.

## 2 Proposed Method

Our approach has two components: (1) Constructing a contextualized instruction-tuning dataset and (2) Fine-tuning foundation models on the built dataset.

### 2.1 Construction of Contextualized Instruction Tuning Dataset

#### Knowledge Tree of MRI entities

Our in-house neurologists and clinicians developed a hierarchical knowledge tree [9] to categorize and annotate entities in MRI reports. The distribution of the major entities is shown in Fig 2, while the hierarchical structure is illustrated in Fig 3. This taxonomy is derived from clinically and systematically validated anatomical locations, lesion types, and imaging features [11, 25]. The tree has three primary nodes: *Element* (shown in blue) – Includes types of lesions, malformations, tumors, and vascular abnormalities; *Location* (shown in red) – Represents the anatomical positioning of elements; and *Other Features* (shown in green) – Covers imaging characteristics such as atrophy, calcification, mass effect and others. The tree has a depth of 4 levels from the root with *Element* being the most complex, comprising four primary subcategories and over 50 distinct subtype elements. *Location* includes three subcategories with 15 distinct positions, while *Other Features* covers 14 distinct imaging attributes. The radial visualization in Fig 3 demonstrates the hierarchical relationships and the breadth of each category, with the full tree available in our code repository **<u>here</u>**.

**Fig. 1.**
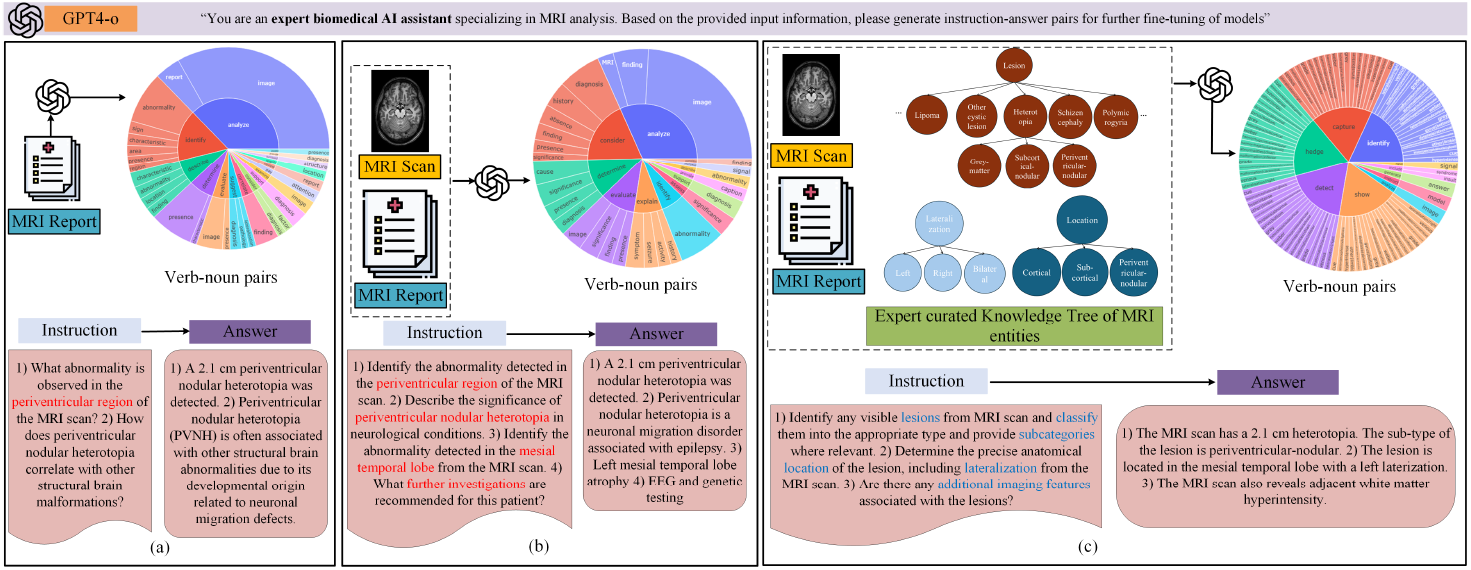
Instruction-tuning approaches: (a) MRI report-based (b) MRI scan and report-based (c) **Proposed TREE-TUNE**: Knowledge Tree with MRI scan and report-based. Includes verb-noun pairs and an instruction-answer sample for each approach.

**Fig. 2.**
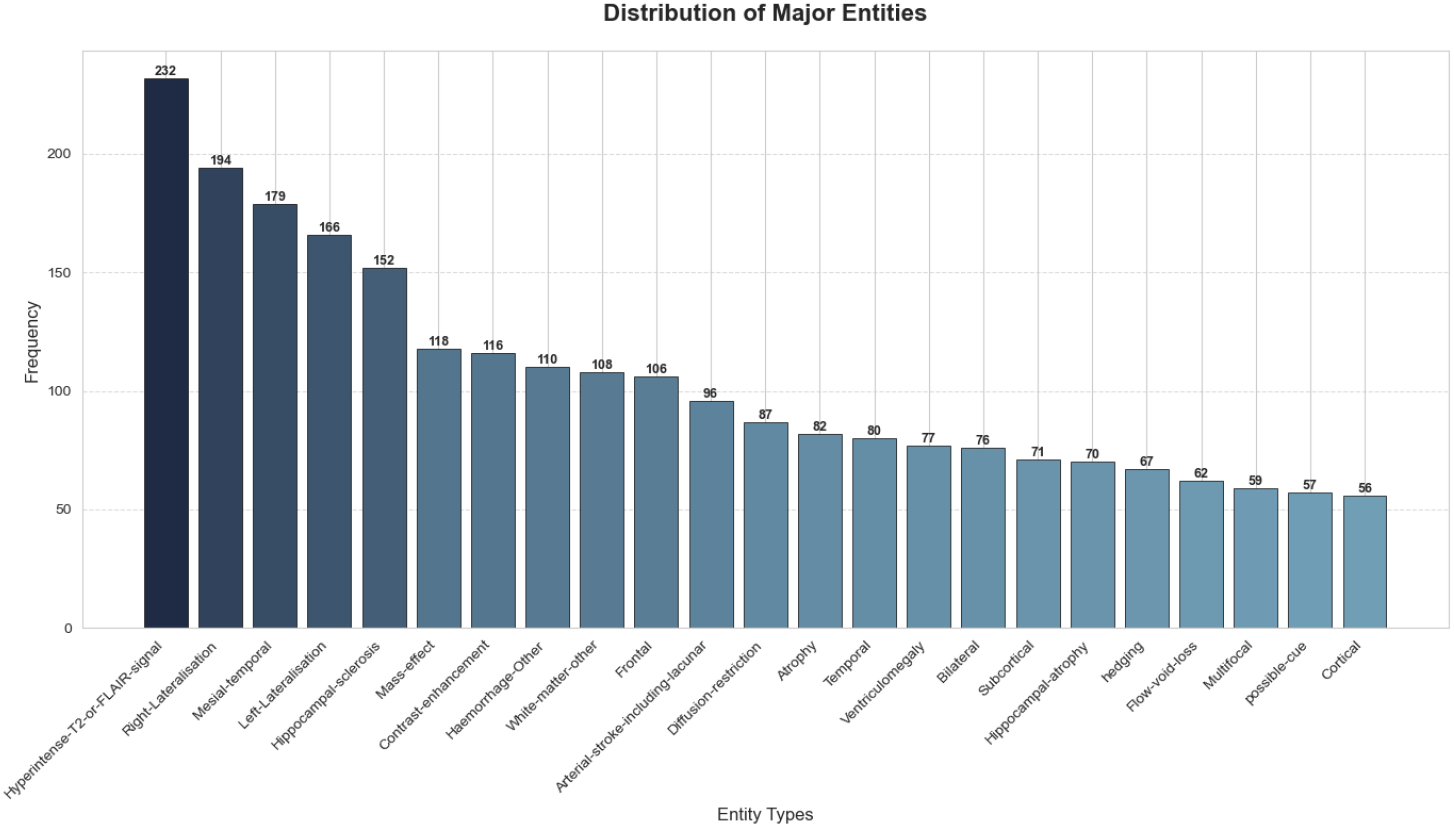
Distribution of Knowledge Tree Major Entities.

**Fig. 3.**
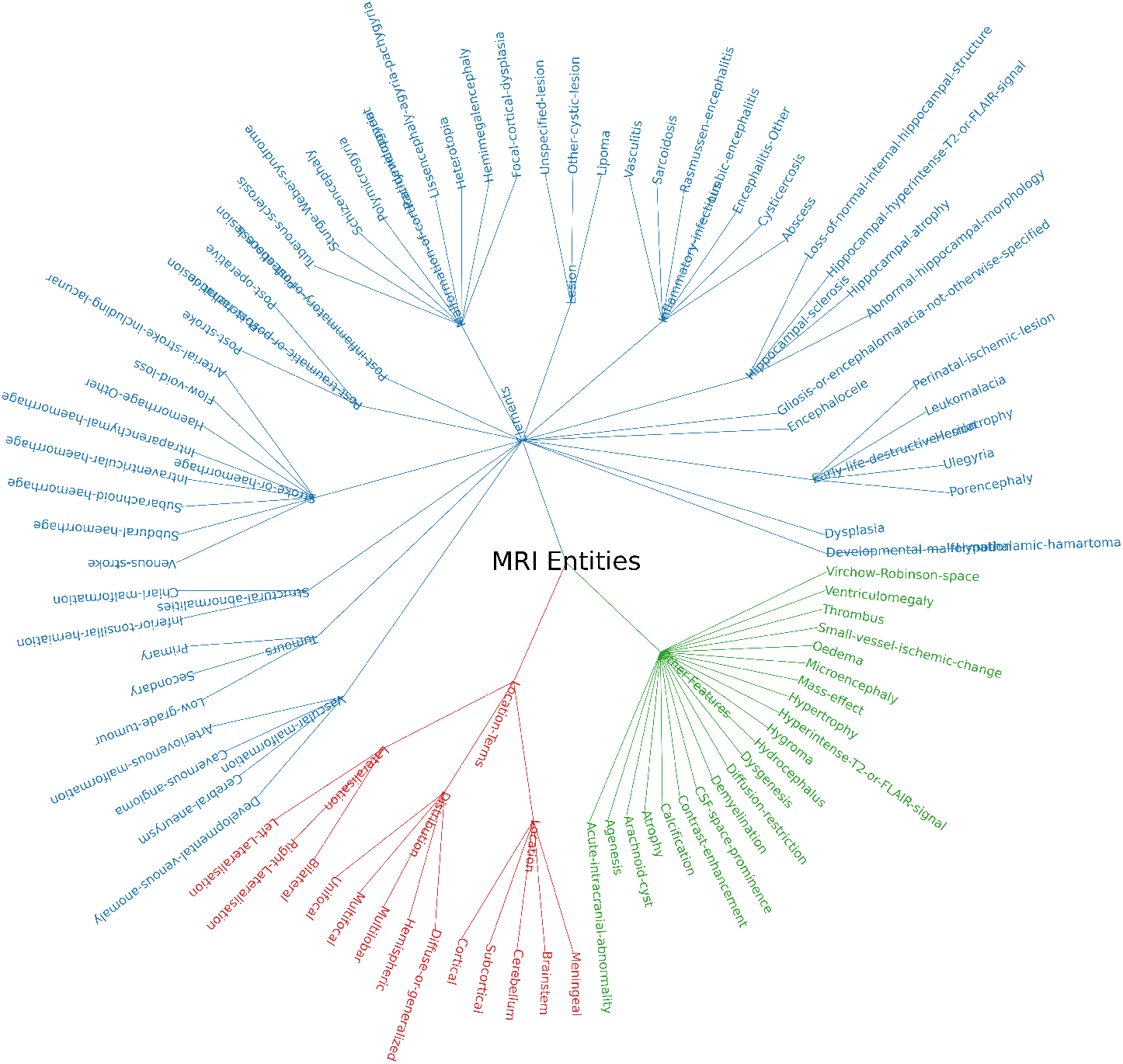
Hierarchical structure of the MRI knowledge tree showing the three main categories: Elements (blue), Location (red), and Other Features (green). The radial layout illustrates the depth and breadth of each category, with terminal nodes representing specific entity types used for annotation.

#### Contextualized Data-specific Generation of Instruction-Answer Pairs

Traditional instruction-answer pair generation follows two strategies: 1) *Text based generation*, where LLMs like GPT-4 generate synthetic pairs from captions or reports [16] (Fig. 1(a)). 2) *Multimodal generation*, where both images and text are provided to multimodal LLMs (e.g., GPT-4v) for enhanced quality [2] (Fig.1(b)). In contrast, our method **TREE-TUNE** (Fig. 1(c)) enhances instruction generation by incorporating three inputs into GPT-4o: an MRI scan, its corresponding report, and the MRI knowledge tree. GPT-4o, prompted (Fig 4) as an “Expert medical AI Assistant specializing in MRI analysis” (temp = 0.1), then generates contextualized instruction-answer (IA) pairs based on observed MRI entities from scan and report and their hierarchical placement in our built knowledge tree. The structured prompt used for this task is shown in Fig. 4, which clearly specifies the inputs and expected outputs in a clinical context to guide consistent IA generation. Our TREE-TUNE generated 4,896 IA pairs across 274 patient cases (median: 24 IA pairs per patient; average length of instruction: 49.1 words).

**Fig. 4.**
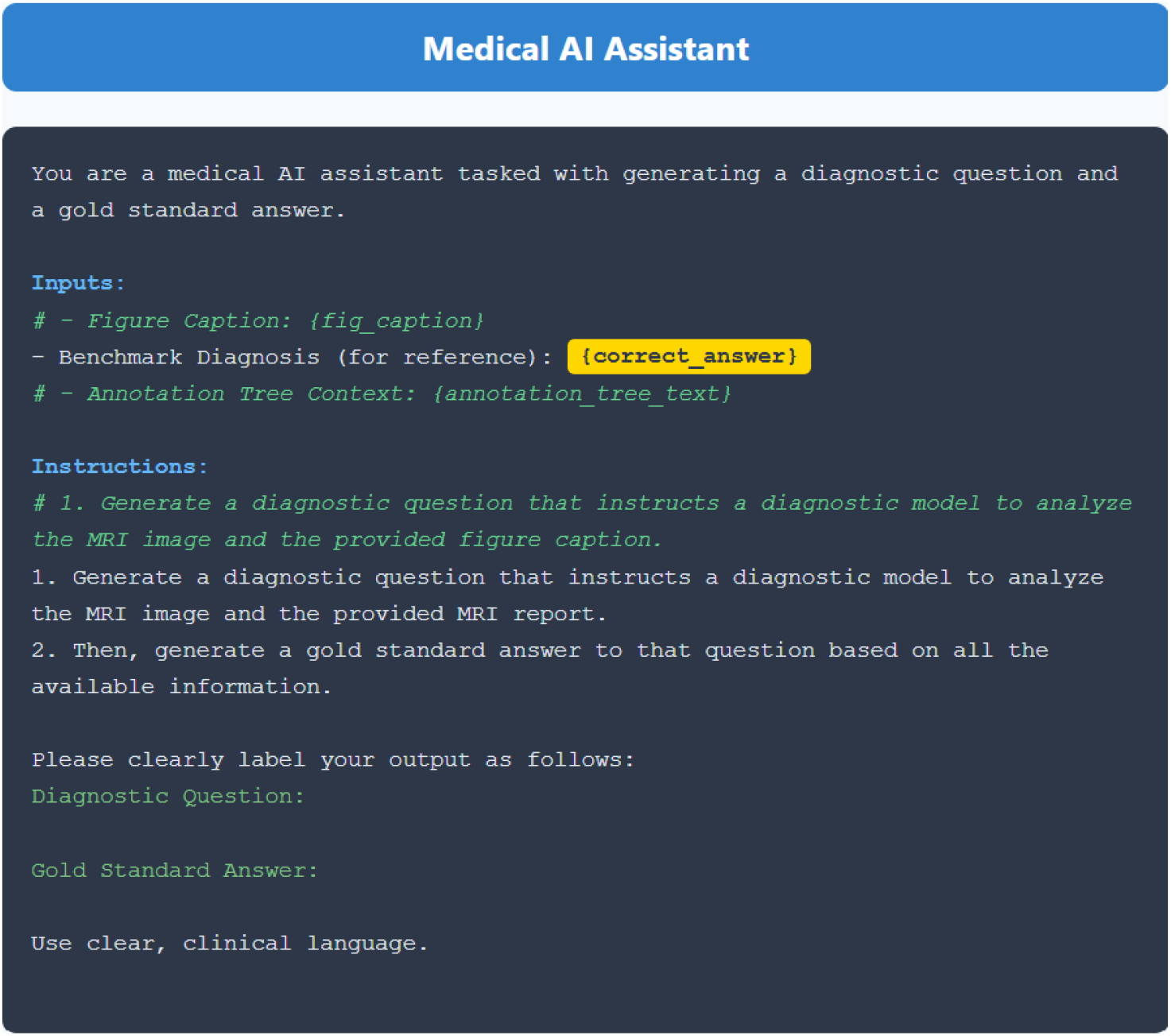
Instruction prompt used in TREE-TUNE. The prompt guides GPT-4o to generate a diagnostic question and a gold standard answer based on an MRI image, MRI report, and knowledge tree context.

For example, if an MRI report mentions “periventricular nodular heterotopia” and “white matter hyperintensity”, the knowledge tree links them to their nodes: *Lesion*→*Heterotopia*→*Periventricular-nodular* and *Other Features* respectively (Fig 1(c)). This enables GPT-4o to generate more nuanced instructions (highlighted in blue in Fig 1(c)), producing more context-aware answers than standard prompting, which embeds terms directly into the instructions (highlighted in red in Fig 1(a),(b)). Similarly, presence of a lesion in the “left mesial temporal lobe” in the MRI scan links it to *Location*→*Lateralization* nodes, resulting in knowledge-informed instructions and more case-specific responses (compare question 3) and question 2) from Fig 1(b) and (c)).

By integrating the knowledge tree with MRI scans and reports, **TREE-TUNE** enhances instruction specificity and contextuality. The verb-noun pair distribution (Fig. 1) shows that our approach yields a richer noun set per verb than standard methods, effectively capturing MRI entities and generating more refined instructions, as further analyzed in the Results section.

### 2.2 Instruction-Tuning of Foundation Models

To evaluate the impact of our contextualized instruction-tuning dataset, we fine-tune foundation models following LLaVA [18] (Fig 5). Given an MRI scan *X*_*I*_, a pretrained biomedical vision encoder extracts visual features *Z*_*I*_ = *g*(*X*_*I*_), which are projected into the language embedding space *H*_*I*_ via a trainable matrix *W*_*I*_. The ground truth ASM, *X*_*ASM*_, is represented in SMILES format [27] and processed as *Z*_*ASM*_ = *d*(*X*_*ASM*_) using the pretrained graph-based drug encoder MoLeR [19], which models molecular structures as graphs and encodes the atoms, bonds, and other structural features of the ASM drug molecule. ASM embeddings *Z*_*ASM*_ are then projected into the language space *H*_*ASM*_ using another trainable matrix *W*_*ASM*_.

**Fig. 5.**
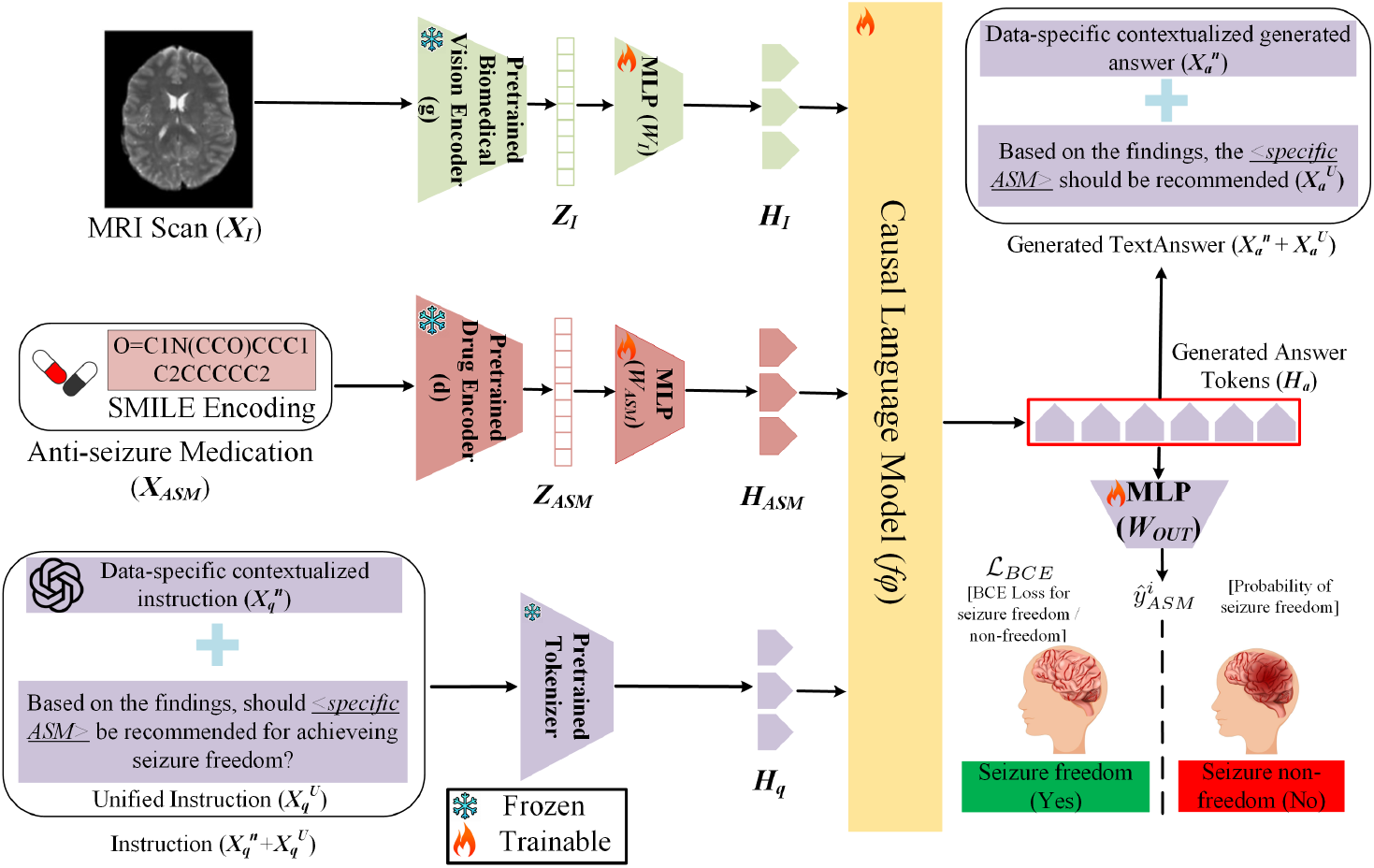
Our overall framework for instruction-tuning of models to generate answer and predict response outcome for an input MRI scan,an ASM drug, and instruction prompt.

For each MRI-ASM pair, we generate a contextualized instruction-answer set using the knowledge tree, MRI scans, and reports. These pairs 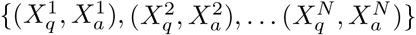serve as ground truth for fine-tuning, where *N* represents the number of pairs per sample. The instruction-answers are structured sequentially, ending with a “unified instruction-answer pair” 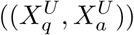 (Fig 5) prompting the binary seizure-free response outcome (Yes/No) for the given ASM. The combined instructions are tokenized into embeddings *H*_*q*_ via a pretrained tokenizer [16] and the language model *f*_*ϕ*_ (Vicuna [7]) is fine-tuned on the generated tokens using its original autoregressive objective. Following [18], for an instruction-answer pair of length *L*, we compute the probability of the target answer *X*_*a*_ by:

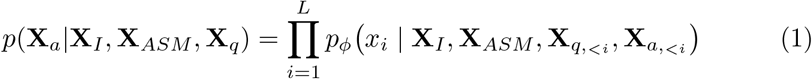

where *ϕ* are trainable parameters of the language model, 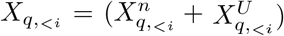, and 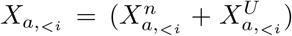 represent the combined *n*^*th*^ instruction-answer pair and the unified instruction-answer pair tokens before the current predicted token *x*_*i*_.

In addition to the autoregressive training objective, we use binary cross entropy loss to optimize the prediction of response outcome for an ASM. As shown in Fig 5, we pass the generated answer tokens *H*_*a*_ to a trainable layer *W*_*OUT*_ followed by a *sigmoid*() function to compute the probability of response outcome:

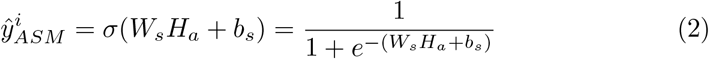

where *W*_*s*_ and *b*_*s*_ are the trainable parameters of *W*_*OUT*_, *H*_*a*_ are the generated answer tokens, and 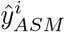 is the predicted probability of seizure freedom/non-freedom for MRI scan *X*_*I*_ and the ASM *X*_*ASM*_. We then apply the standard BCE loss between the predicted outcome probability and the ground truth [1,0] corresponding to seizure freedom / non-freedom.

Our framework uses direct fine-tuning, freezing the pretrained biomedical vision encoder *g*_*θ*_ and drug encoder *d*_*ω*_, while updating only the projection matrices *W*_*I*_ (image), *W*_*ASM*_ (drug), *W*_*OUT*_ (outcome) and the language model *f*_*ϕ*_. We optimize the network using both autoregressive and BCE loss with equal weighting. During inference, we only instruct the framework with the unified instruction 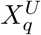 and obtain the generated answer and the outcome response in the manner described earlier.

### 2.3 Training & Implementation Details

To construct the instruction-tuning datasets, we used the GPT-4o API (Jan–Feb 2025). No data augmentation was applied to MRI scans. Fine-tuning started with a learning rate of 1 *×* 10^*−*4^, using a cosine scheduler, 5-epoch warm-up, and a constant schedule type. Training ran for 100 epochs with a batch size of 16 per device using the AdamW optimizer. Following prior ASM recommendation studies [12], we evaluated performance using AUC, precision, and recall. Additionally, semantic similarity matching assessed the fine-tuned language model’s responses. Experiments were conducted on NVIDIA A100 and A6000 GPUs.

## 3 Datasets and Splits

We retrospectively collected two private datasets: **ASM-ED1**, which is available upon request for research purposes only, and **ASM-ED2**, which could be curated from the Human Epilepsy Project [10]. Both datasets originate from distinct cohorts of newly diagnosed epilepsy patients. Seizure control was assessed one year after starting the first ASM, with success defined as seizure freedom while maintaining the same ASM.

### ASM Distribution

Patients were administered with one of the seven ASMs: carbamazepine, lamotrigine, levetiracetam, oxcarbazepine, phenytoin, topiramate, or valproate. The most frequently used were levetiracetam, lamotrigine, oxcarbazepine, and carbamazepine, having 250, 79, 37, and 24 cases in ASM-ED1, and 116, 41, 39, and 25 cases in ASM-ED2. The remaining three ASMs had 54 cases only in ASM-ED1. ASM-ED1 included 444 patients (112 seizure-free, 332 not), while ASM-ED2 had 247 patients (62 seizure-free, 185 not).

### Train and Test splits

We use 5-fold cross-validation with an 80/20 train-validation split for both datasets. ASM-ED1 is used to evaluate unseen ASM recommendation, as it includes all seven ASMs. Training is limited to the four most common ASMs (levetiracetam, lamotrigine, oxcarbazepine, and carbamazepine), while valproate, topiramate, and phenytoin are encountered only in testing.

## 4 Results

We analyze the lexical composition of our instruction-tuning dataset and also benchmark various biomedical foundation models (vision and vision-language) alongside different instruction-tuning approaches for ASM outcome prediction.

### 4.1 Lexical Analysis of Instruction-Tuning Approaches

We analyzed the lexical composition of instructions from different generation approaches (Table 1). TREE-TUNE shows the richest set of nouns and adjectives, significantly surpassing MRI report- or scan-based methods. Its significantly higher noun percentage captures more epilepsy-specific entities (e.g., lesion-encephalocele-gliosis, lateralization-right), while significant proportion of adjectives enhance descriptive detail, improving characterization of epilepsy-related MRI findings, as also shown in the verb-noun pair chart (Fig. 1).

**Table 1.** Comparison of lexical composition (in terms of %) across all instructions for the different instruction tuning approaches on both datasets combined.

| Dataset / Lexical | noun | adjective | adverb | verb | numerical | preposition |
| --- | --- | --- | --- | --- | --- | --- |
| MRI report [16] | 34.1 | 16.3 | 1.5 | 12.9 | 0.7 | 11.9 |
| MRI scan + report [2] | 36.9 | 12.3 | 0.8 | 15.6 | 0.1 | 9.4 |
| **TREE-TUNE (Ours)** | **39.9** | **20.9** | 1.6 | 15.6 | 0.7 | 3.6 |

### 4.2 ASM Outcome Prediction Performance

We evaluate ASM outcome prediction in two settings: **seen ASMs** (trained) and **zero-shot unseen ASMs** (not seen during training). Table 2 shows seen ASM results, comparing vision-only, vision-language, and instruction-tuning methods, including the naïve prompt “Describe the findings in the MRI scan”. Instruction-tuning consistently outperforms other methods, with our approach achieving the highest AUC (76.45 (p=0.012)) on ASM-ED1, 63.03 (p=0.041) on unseen ASMs, showing a **5.53**% average AUC gain over standard MRI report-based methods. ASM-ED2 has fewer samples and greater hospital variability, while ASM-ED1 (from only two hospitals) performs better. The challenging ASM-ED2 and unseen ASM settings highlight TREE-TUNE’s strong performance. Fig. 6 illustrates ROC curves of AUC scores for four commonly prescribed ASMs, showing that OXC and CBZ achieve the highest predictive performance.

**Table 2.** Seizure outcome prediction performance for popular seen ASMs.

| Method / Dataset / Metric | ASM-ED1 |  |  | ASM-ED2 |  |  |
| --- | --- | --- | --- | --- | --- | --- |
|  | AUC (%) | Precision (%) | Recall (%) | AUC (%) | Precision (%) | Recall (%) |
| BiomedCLIP Vision [30] | 59.25<sub>1.15</sub> | 48.56<sub>1.12</sub> | 51.20<sub>1.14</sub> | 55.53<sub>1.30</sub> | 46.01<sub>1.20</sub> | 49.50<sub>1.30</sub> |
| LLaVA-Med Vision [16] | 60.37<sub>1.12</sub> | 49.58<sub>3.11</sub> | 52.43<sub>2.13</sub> | 58.91<sub>1.25</sub> | 48.21<sub>3.02</sub> | 51.74<sub>2.21</sub> |
| BiomedCLIP Vision-Language [30] | 65.00<sub>1.90</sub> | 54.02<sub>1.73</sub> | 53.01<sub>2.32</sub> | 51.86<sub>3.72</sub> | 52.00<sub>2.20</sub> | 51.01<sub>2.50</sub> |
| **Instruction-Tuning Approaches** |  |  |  |  |  |  |
| Naive | 74.71<sub>2.13</sub> | 69.63<sub>5.15</sub> | 63.59<sub>1.24</sub> | 61.36<sub>3.65</sub> | 67.03<sub>5.53</sub> | 62.31<sub>1.65</sub> |
| MRI report [16] | 71.54<sub>4.35</sub> | 68.23<sub>4.76</sub> | 62.87<sub>2.83</sub> | 60.21<sub>5.12</sub> | 58.44<sub>5.97</sub> | 52.23<sub>3.44</sub> |
| MRI scan + report [2] | 75.24<sub>5.81</sub> | 69.57<sub>4.86</sub> | 65.76<sub>4.32</sub> | 61.75<sub>4.87</sub> | 61.86<sub>1.16</sub> | 60.24<sub>3.91</sub> |
| **TREE-TUNE (Ours)** | **76.45<sub>4.21</sub>** | **70.52<sub>6.12</sub>** | **67.85<sub>6.57</sub>** | **66.33<sub>6.15</sub>** | **61.94<sub>4.92</sub>** | **55.42<sub>6.92</sub>** |
| Average Increment | 4.94 | 2.29 | 4.98 | 6.12 | 3.5 | 3.19 |

**Fig. 6.**
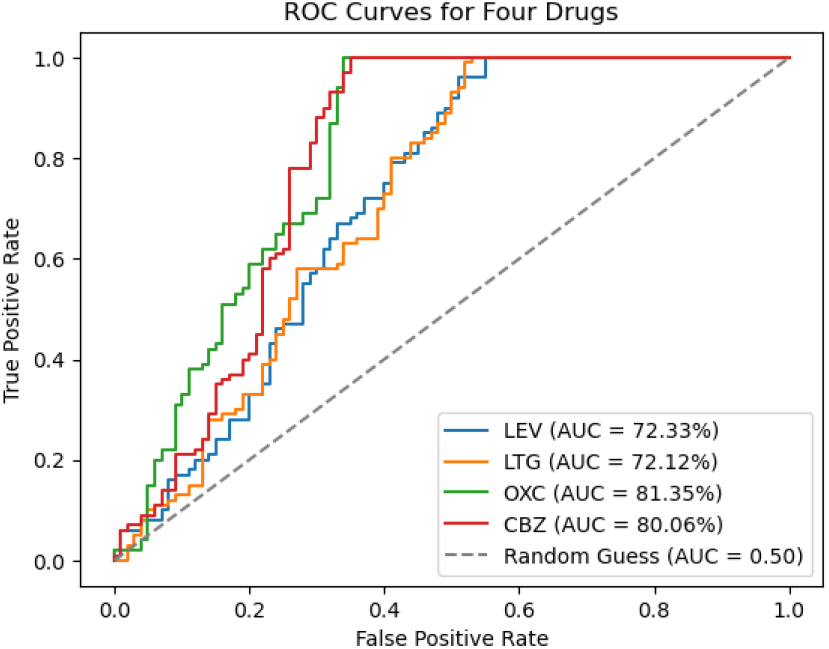
ROC curves and AUC scores for four ASMs (LEV, LTG, OXC, CBZ). Our model shows higher capability in predicting seizure outcomes for the drugs OXC and CBZ.

#### Zero-shot prediction

We evaluate TREE-TUNE’s zero-shot capability on three unseen ASMs excluded from ASM-ED1 training. As shown in Table 3, our method improves AUC by **3.51%** over MRI report-based instruction tuning, outperforming others and maintaining robust generalization.

**Table 3.** Seizure outcome prediction performance for unseen ASMs.

| Method / Metric | AUC (%) | Precision (%) | Recall (%) |
| --- | --- | --- | --- |
| BiomedCLIP Vision [30] | 55.01 | 35.03 | 30.14 |
| BiomedCLIP Vision-Language [30] | 58.05 | 40.04 | 39.07 |
| **Instruction-tuning approaches** |  |  |  |
| Naive | 60.12 | 45.06 | 48.68 |
| MRI report [16] | 59.52 | 51.03 | 59.05 |
| MRI scan + report [2] | 60.07 | 56.05 | 61.04 |
| **TREE-TUNE (Ours)** | **63.03** | **60.06** | **65.07** |
| Average Increment | 3.51 | 9.03 | 6.02 |

### 4.3 Qualitative Analysis of TREE-TUNED vs. Standard Models

Fig 7 compares TREE-TUNE, GPT-4o, and LLaVA-Med responses to a zero-shot prompt and MRI scan. GPT-4o and LLaVA-Med mislabel findings of cortical dysplasia, juxtacortical white matter abnormalities, and gray-white junction blurring as hippocampal sclerosis and mesial temporal lobe epilepsy. In contrast, TREE-TUNE accurately identifies these features, achieves the highest semantic similarity, and predicts ASM probability aligned with the ground truth. This demonstrates superior anatomical understanding and a step towards reasoning.

**Fig. 7.**
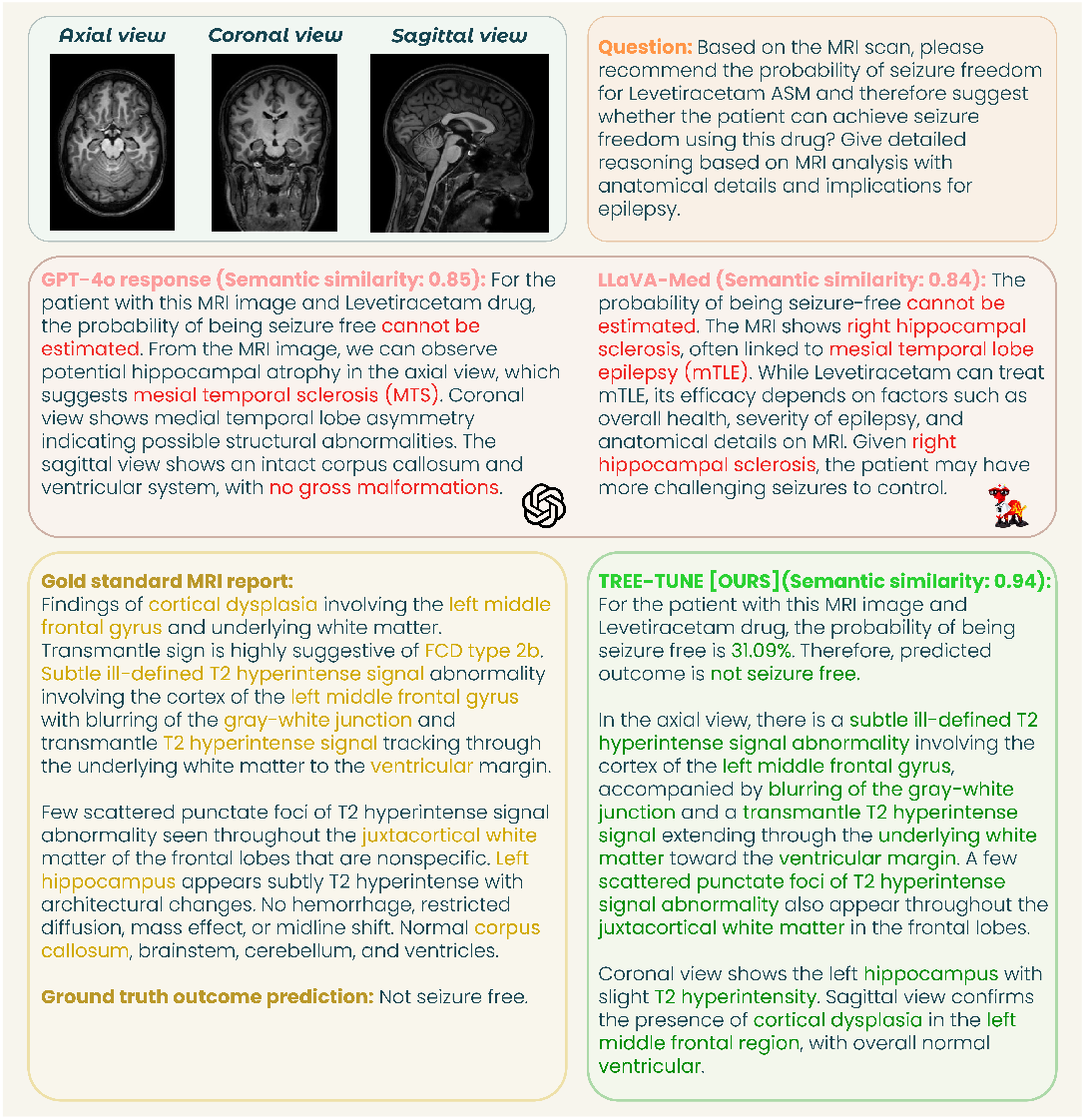
An exemplar case to demonstrate the generated response of our TREE-TUNE.

## 5 Conclusion

This work evaluates the effectiveness of biomedical foundation models in predicting ASM outcomes from MRI scans and reports. We introduce TREE-TUNE, a novel MRI knowledge tree driven contextualized instruction-tuning framework to enhance model performance beyond standard instruction-tuning strategies. Moreover, our approach not only achieves high accuracy in recommending commonly prescribed ASMs but also gives reasonable performance for unseen ASMs. Our work marks a foundational step toward developing a reasoning-based ASM recommendation system, paving the way for personalized epilepsy management.

## Data Availability

All data produced in the present study are available upon reasonable request to the authors

## Notes

### Competing Interest Statement

The authors have declared no competing interest.

### Funding Statement

This study did not receive any funding

### Author Declarations

Alfred Ethics Committee - granted.

## References

1. Bernasconi, A., Cendes, F., Theodore, W.H., Gill, R.S., Koepp, M.J., Hogan, R.E., Jackson, G.D., Federico, P., Labate, A., Vaudano, A.E., et al.: Recommendations for the use of structural magnetic resonance imaging in the care of patients with epilepsy: a consensus report from the international league against epilepsy neuroimaging task force. Epilepsia 60(6), 1054–1068 (2019)

2. Chen, L., Li, J., Dong, X., Zhang, P., He, C., Wang, J., Zhao, F., Lin, D.: Sharegpt4v: Improving large multi-modal models with better captions. In: European Conference on Computer Vision. pp. 370–387. Springer (2024)

3. Chen, Z., Brodie, M.J., Kwan, P.: What has been the impact of new drug treatments on epilepsy? Current opinion in neurology 33(2), 185–190 (2020)

4. Chen, Z., Brodie, M.J., Liew, D., Kwan, P.: Treatment outcomes in patients with newly diagnosed epilepsy treated with established and new antiepileptic drugs: a 30-year longitudinal cohort study. JAMA neurology 75(3), 279–286 (2018)

5. Chen, Z., Rollo, B., Antonic-Baker, A., Anderson, A., Ma, Y., O’Brien, T.J., Ge, Z., Wang, X., Kwan, P.: Brain health: New era of personalised epilepsy management. The BMJ 371 (2020)

6. Cheval, M., Houot, M., Chastan, N., Szurhaj, W., Marchal, C., Catenoix, H., Valton, L., Gavaret, M., Herlin, B., Biraben, A., et al.: Early identification of seizure freedom with medical treatment in patients with mesial temporal lobe epilepsy and hippocampal sclerosis. Journal of Neurology 270(5), 2715–2723 (2023)

7. Chiang, W.L., Li, Z., Lin, Z., Sheng, Y., Wu, Z., Zhang, H., Zheng, L., Zhuang, S., Zhuang, Y., Gonzalez, J.E., et al.: Vicuna: An open-source chatbot impressing gpt-4 with 90%* chatgpt quality. See https://vicuna.lmsys.org (accessed 14 April 2023) 2(3), 6 (2023)

8. Croce, P., Ricci, L., Pulitano, P., Boscarino, M., Zappasodi, F., Lanzone, J., Narducci, F., Mecarelli, O., Di Lazzaro, V., Tombini, M., et al.: Machine learning for predicting levetiracetam treatment response in temporal lobe epilepsy. Clinical Neurophysiology 132(12), 3035–3042 (2021)

9. Foster, E., Fazio, T., Holper, S., Truong, T., Verspoor, K.: Melbourne brain mri report epilepsy annotation dataset (aug 2025). 10.17632/6kxnvb7yyg.1

10. Fox, J., Barnard, S., Agashe, S.H., Holmes, M.G., Gidal, B., Klein, P., AbouKhalil, B.W., French, J., Investigators, H.E.P.: Patterns of antiseizure medication utilization in the human epilepsy project. Epilepsia 64(12), 3196–3204 (2023)

11. Hakami, T., Mcintosh, A., Todaro, M., Lui, E., Yerra, R., Tan, K.M., French, C., Li, S., Desmond, P., Matkovic, Z., et al.: Mri-identified pathology in adults with new-onset seizures. Neurology 81(10), 920–927 (2013)

12. Hakeem, H., Feng, W., Chen, Z., Choong, J., Brodie, M.J., Fong, S.L., Lim, K.S., Wu, J., Wang, X., Lawn, N., et al.: Development and validation of a deep learning model for predicting treatment response in patients with newly diagnosed epilepsy. JAMA neurology 79(10), 986–996 (2022)

13. Hu, Z., Jiang, D., Zhao, X., Yang, J., Liang, D., Wang, H., Zhao, C., Liao, J.: Predicting drug treatment outcomes in children with tuberous sclerosis complex– related epilepsy: A clinical radiomics study. American Journal of Neuroradiology 44(7), 853–860 (2023)

14. Labate, A., Aguglia, U., Tripepi, G., Mumoli, L., Ferlazzo, E., Baggetta, R., Quattrone, A., Gambardella, A.: Long-term outcome of mild mesial temporal lobe epilepsy: a prospective longitudinal cohort study. Neurology 86(20), 1904–1910 (2016)

15. Lee, H.M., Fadaie, F., Gill, R., Caldairou, B., Sziklas, V., Crane, J., Hong, S.J., Bernhardt, B.C., Bernasconi, A., Bernasconi, N.: Decomposing mri phenotypic heterogeneity in epilepsy: a step towards personalized classification. Brain 145(3), 897–908 (2022)

16. Li, C., Wong, C., Zhang, S., Usuyama, N., Liu, H., Yang, J., Naumann, T., Poon, H., Gao, J.: Llava-med: Training a large language-and-vision assistant for biomedicine in one day. Advances in Neural Information Processing Systems 36 (2024)

17. Liu, H., Li, C., Li, Y., Lee, Y.J.: Improved baselines with visual instruction tuning. In: Proceedings of the IEEE/CVF Conference on Computer Vision and Pattern Recognition. pp. 26296–26306 (2024)

18. Liu, H., Li, C., Wu, Q., Lee, Y.J.: Visual instruction tuning. Advances in neural information processing systems 36, 34892–34916 (2023)

19. Maziarz, K., Jackson-Flux, H., Cameron, P., Sirockin, F., Schneider, N., Stiefl, N., Segler, M., Brockschmidt, M.: Learning to extend molecular scaffolds with structural motifs. arXiv preprint 2103.03864 (2021)

20. Perucca, E., Brodie, M.J., Kwan, P., Tomson, T.: 30 years of second-generation antiseizure medications: impact and future perspectives. The Lancet Neurology 19(6), 544–556 (2020)

21. Sepehri, M.S., Fabian, Z., Soltanolkotabi, M., Soltanolkotabi, M.: Mediconfusion: Can you trust your ai radiologist? probing the reliability of multimodal medical foundation models. arXiv preprint 2409.15477 (2024)

22. Shazadi, K., Petrovski, S., Roten, A., Miller, H., Huggins, R.M., Brodie, M.J., Pirmohamed, M., Johnson, M.R., Marson, A.G., O’Brien, T.J., et al.: Validation of a multigenic model to predict seizure control in newly treated epilepsy. Epilepsy research 108(10), 1797–1805 (2014)

23. Shin, Y., Hwang, S., Lee, S.B., Son, H., Chu, K., Jung, K.Y., Lee, S.K., Park, K.I., Kim, Y.G.: Using spectral and temporal filters with eeg signal to predict the temporal lobe epilepsy outcome after antiseizure medication via machine learning. Scientific Reports 13(1), 22532 (2023)

24. Silva-Alves, M.S., Secolin, R., Carvalho, B.S., Yasuda, C.L., Bilevicius, E., Alvim, M.K., Santos, R.O., Maurer-Morelli, C.V., Cendes, F., Lopes-Cendes, I.: A prediction algorithm for drug response in patients with mesial temporal lobe epilepsy based on clinical and genetic information. PLoS One 12(1), e0169214 (2017)

25. Truong, T.H., Foster, E., Fazio, T., Holper, S., Verspoor, K.M.: Exploring the limits of llms in low-resource information extraction: Case study in brain mri reports for epilepsy. medRxiv (2025). 10.1101/2025.08.02.25332570, https://www.medrxiv.org/content/early/2025/08/05/2025.08.02.25332570

26. Wang, X., Hu, T., Yang, Q., Jiao, D., Yan, Y., Liu, L.: Graph-theory based degree centrality combined with machine learning algorithms can predict response to treatment with antiepileptic medications in children with epilepsy. Journal of Clinical Neuroscience 91, 276–282 (2021)

27. Weininger, D.: Smiles, a chemical language and information system. 1. introduction to methodology and encoding rules. Journal of chemical information and computer sciences 28(1), 31–36 (1988)

28. World Health Organization: Epilepsy (2024), https://www.who.int/news-room/fact-sheets/detail/epilepsy, accessed: 13 Feb. 2025

29. Xiong, Z., Wang, X., Zhou, Y., Keane, P.A., Tham, Y.C., Wang, Y.X., Wong, T.Y.: How generalizable are foundation models when applied to different demographic groups and settings? NEJM AI 2(1), AIcs2400497 (2025)

30. Zhang, S., Xu, Y., Usuyama, N., Xu, H., Bagga, J., Tinn, R., Preston, S., Rao, R., Wei, M., Valluri, N., et al.: Biomedclip: a multimodal biomedical foundation model pretrained from fifteen million scientific image-text pairs. arXiv preprint 2303.00915 (2023)

